# Preoperative μ-opioid receptor availability predicts weight loss following bariatric surgery

**DOI:** 10.1101/2021.01.27.21250121

**Authors:** Henry K. Karlsson, Lauri Tuominen, Semi Helin, Paulina Salminen, Pirjo Nuutila, Lauri Nummenmaa

**Affiliations:** Turku PET Centre, University of Turku, Turku, Finland; Institute of Mental Health Research, University of Ottawa, Ottawa, Ontario, Canada; Department of Digestive Surgery, University of Turku and Turku University Hospital, Turku, Finland; Department of Endocrinology, Turku University Hospital, Turku, Finland; Department of Psychology, University of Turku, Turku, Finland

**Keywords:** Obesity, positron emission tomography, reward, opioid, dopamine, receptors, bariatric surgery, weight loss

## Abstract

**Background:** Bariatric surgery is the most effective method for weight loss in morbid obesity. There is significant individual variability in the weight loss outcomes, yet factors leading to postoperative weight loss or weight regain remain elusive. Alterations in the µ-opioid receptor (MOR) and dopamine D_2_ receptor (D_2_R) systems are associated with obesity, appetite control, and reward processing. The magnitude of initial brain receptor system perturbation is a plausible predictor of long-term surgical weight loss outcomes. The aim was to test this hypothesis by measuring obese subjects’ MOR and D_2_R availability with positron emission tomography (PET) preoperatively before bariatric surgery and then assessing their weight development association with regional MOR and D_2_R availabilities at 2-year follow-up.

**Methods:** We studied 19 morbidly obese women (mean BMI 40, mean age 43) scheduled to undergo bariatric surgery, i.e. Roux-en-Y gastric bypass or sleeve gastrectomy, according to their standard clinical treatment. Preoperative MOR and D_2_R availabilities were measured using PET with [^11^C]carfentanil and [^11^C]raclopride, respectively. Subject weight was recorded at 3, 6, 12, and 24 months after surgery. Radiotracer binding potentials (*BP*_ND_) were extracted and correlated with patient weight at different time points. ROIs were delineated in the striatum and in limbic and paralimbic components of the emotion and reward networks.

**Results:** MOR availabilities were not correlated with preoperative weight. MOR availabilities in the amygdala (*r* = −0.54), insula (*r* = −0.46), ventral striatum (*r* = −0.48) and putamen (*r* = −0.49) were associated with subject weight at 3 months. Significant association was found in the amygdala at 6 months (*r* = −0.53), 12 (*r* = −0.49), and 24 months (*r* = −0.50). D_2_R availabilities were associated with neither preoperative weight nor weight loss at any follow-up time point.

**Conclusions:** To our knowledge, this is the first study to demonstrate that neuroreceptor markers prior to bariatric surgery in patients with morbid obesity are associated with the postoperative weight loss. Preoperative MOR availability in the amygdala was associated with long-term postoperative weight development after surgery suggesting that postoperative weight regain may derive from dysfunction in the opioid system. Postoperative weight loss outcomes after bariatric surgery may be partially predicted based on preoperative receptor availability opening up new potential for treatment possibilities.

**Clinical Trials Registration:** SleevePET2, NCT01373892, http://www.clinicaltrials.gov

## Introduction

The prevalence of obesity is constantly increasing and reaching global pandemic levels. Accumulating evidence suggests that dysfunctions in appetite control and reward processing mechanism significantly contribute to weight gain and maintenance, and particularly brain’s dopamine and opioid systems in the reward circuit are dysfunctional in obesity. Dopamine D_2_receptor (D_2_R) expression and function are altered in obesity (Volkow et al., 2008; Stice et al., 2011; Salamone and Correa, 2013), whereas endogenous opioid system is consistently linked to hedonic aspects of feeding in animals (Pecina and Smith, 2010; Fields and Margolis, 2015). In humans, feeding triggers endogenous opioid release (Tuulari et al., 2017) and accordingly pharmacological challenge studies have found that both µ-opioid receptor (MOR) antagonists and inverse agonists reduce human eating behaviour (Nathan et al., 2012; Cambridge et al., 2013). MOR levels are also downregulated in obese subjects, underlining the importance the opioid system perturbation in overeating (Burghardt et al., 2015; Karlsson et al., 2015).

Bariatric surgery is currently the most effective method for weight loss in obesity. Mean postoperative total weight loss of 27 % has been shown among patients even after 12 years (Adams et al., 2017). Bariatric surgery procedures are also much more effective than intensive medical therapy to reach glycemic control (Schauer et al., 2017). For weight loss, there is currently some consensus to use standardized reporting guidelines in bariatric surgery literature (Brethauer et al., 2015), but similar uniform consensus needs to be achieved regarding postoperative weight regain in order to assess the durability of weight loss and to reliably evaluate potential treatment options (Salminen, 2018). Weight regain following bariatric surgery occurs in one fifth (Shantavasinkul et al., 2016; Courcoulas et al., 2018; Baig et al., 2019) up to one third of the patients (Cooper et al., 2015; Monaco-Ferreira and Leandro-Merhi, 2017; Voorwinde et al., 2020).

Factors leading to weight regain following surgery remain poorly understood, yet cross-sectional studies point towards a possible role of the brain in regulating the treatment response. Impulsivity and disinhibition are traits often associated with poorer weight loss after surgery, but both psychosocial issues and psychiatric comorbidities may also have a major impact on weight loss outcomes (Odom et al., 2010; Mauro et al., 2019; Muller et al., 2019; Sarwer et al., 2019). However, only few neuroimaging studies have examined neural predictors of weight loss after surgery. To our knowledge, there are only two small MRI studies that have investigated brain markers that might affect the weight loss outcome of the surgery. Functional connectivity and alterations in brain activity in some of the key areas of reward circuit predicted weight loss 12 months after sleeve gastrectomy (Holsen et al., 2018; Cerit et al., 2019). However, the role of specific neurotransmitter systems – such as D_2_R and MOR implicated in feeding and reward processing – on post-surgical weigh gain and loss remain unknown. In the present study, we addressed this issue by measuring obese subjects’ MOR and D_2_R availability with positron emission tomography (PET) before they underwent bariatric surgery. We followed the subjects for two years and predicted their weight loss outcomes with regional MOR and D_2_R availabilities. We show that MOR availability particularly in the amygdala predicts long-term outcome of the bariatric surgery, suggesting a causal role of this region in appetite control and food intake.

## Methods

The study was conducted in accordance with the Declaration of Helsinki and approved by the Ethical Committee of the Hospital District of South-Western Finland (SleevePET2, NCT01373892, http://www.clinicaltrials.gov). All participants signed ethical committee-approved, informed consent form prior to scans. Group differences in receptor availabilities between normal-weight and morbidly obese subjects have been previously reported for a subset of the subjects (Karlsson et al., 2015; Karlsson et al., 2016).

### Participants

We studied 19 morbidly obese women (mean BMI 40, mean age 43) scheduled to undergo bariatric surgery, i.e. Roux-en-Y gastric bypass or sleeve gastrectomy, according to their standard clinical treatment. Subject characteristics are shown in the Table 1. Clinical screening of the subjects included history, physical examination, anthropometric measurements, and laboratory tests. Exclusion criteria for this study involved opiate drug use, neurological and severe mental disorders, substance abuse, and excessive alcohol consumption determined by clinical interviews, medical history, and blood tests. Seven subjects were smokers (3-15 cigarettes per day). Antidiabetic, antihypertensive and cholesterol lowering drugs were paused prior to the study. Subject weight was recorded before surgery as well as at 3, 6, 12, and 24 months after surgery during a standard hospital visit. Two subjects dropped out of the study before 24 months follow-up visit, but their weight data at 3, 6, and 12 months were included in the analysis. Baseline depressive and anxiety symptoms were recorded using Beck Depression Index II (BDI-II) and State-Trait Anxiety Inventory (STAI), respectively (Spielberger et al., 1983; Beck et al., 1996).

**Table 1:** Characteristics of the participants (*N* = 19). Data are presented as mean ± SD.

|  |  |
| --- | --- |
| Age (y) | 43.3 ± 8.2 |
| Preoperative weight (kg) | 109.8 ± 12,8 |
| Height (cm) | 164.5 ± 5.0 |
| BMI (kg/m<sup>2</sup>) | 40.4 ± 4.1 |
| Amount of alcohol use (units per week) | 1.5 ± 1.7 |
| Tobacco smokers / non-smokers ($N$) | 7 / 12 |
| BDI-II score | 5.4 ± 5.5 |
| STAI score (Trait anxiety) | 37.7 ± 8.1 |
| Injected activity of [<sup>11</sup>C]carfentanil (MBq) | 252.2 ± 10.8 |
| Injected activity of [<sup>11</sup>C]raclopride (MBq) | 248.4 ± 21.9 |

### Image acquisition, quantification of receptor availability, and statistical analysis

We measured µ-opioid receptor availability with the high-affinity agonist [^11^C]carfentanil (Frost et al., 1985) and D_2_ receptor availability with the antagonist [^11^C]raclopride (Farde et al., 1986) using positron emission tomography (PET). Brain scans were performed before the start of the standard very low-calorie diet. Radiotracer production has been described previously (Karlsson et al., 2015). [^11^C]carfentanil and [^11^C]raclopride scans were performed on separate days. Both radiotracers had high radiochemical purity (>99 %). Before scanning, a catheter was placed in the subject’s left antecubital vein for tracer administration. Head was strapped to the scanner table in order to prevent head movement. Subjects fasted two hours prior to scanning. A CT scan was performed to serve as attenuation map. Clinical well-being of subjects were monitored during the scanning.

We injected both tracers as bolus (252.2 ± 10.8 MBq [^11^C]carfentanil and 248.4 ± 21.9 MBq [^11^C]raclopride). After the injection, radioactivity in brain was measured with the GE Heatlhcare Discovery ™ 690 PET/CT scanner (General Electric Medical Systems, Milwaukee, WI, USA) for 51 minutes, using 13 time frames. MR imaging was performed with Philips Gyroscan Intera 1.5 T CV Nova Dual scanner to exclude structural abnormalities and to provide anatomical reference images for the PET scans. High-resolution anatomical images (1 mm^3^voxel size) were acquired using a T1-weighted sequence (TR 25 ms, TE 4.6 ms, flip angle 30°, scan time 376 s).

All alignment and coregistration steps were performed using SPM8 software (www.fil.ion.ucl.ac.uk/spm/) running on Matlab R2012a (The Mathworks Inc., Sherborn, Massachusetts). To correct for head motion, dynamic PET images were first realigned frame-to-frame. The individual T1-weighted MR images were coregistered to the summation images calculated from the realigned frames. Regions of interest (ROIs) for reference regions were drawn manually on MRI images using PMOD 3.4 software (PMOD Technologies Ltd., Zurich, Switzerland). Occipital cortex was used as the reference region for [^11^C]carfentanil and cerebellum for [^11^C]raclopride. Receptor availability was expressed in terms of *BP*_ND_, which is the ratio of specific to non-displaceable binding in brain. *BP*_ND_ was calculated applying basis function method for each voxel using the simplified reference tissue model (SRTM) with reference tissue time activity curves (TAC) as input data (Gunn et al., 1997).

The subject-wise parametric *BP*_ND_ images were normalized to the MNI space using the T1-weighted MR images, and smoothed with a Gaussian kernel of 8 mm FWHM. Anatomic regions of interest were generated in ventral striatum, dorsal caudate nucleus, and putamen using the AAL (Tzourio-Mazoyer et al., 2002) and Anatomy (Eickhoff et al., 2005) toolboxes. Regional [^11^C]carfentanil and [^11^C]raclopride binding potentials (*BP*_ND_) were extracted and correlated with subject weights at 3, 6, 12, and 24 months after surgery. Moreover, BDI and STAI scores were correlated with [^11^C]carfentanil and [^11^C]raclopride binding potentials as well as subject weight at different time points.

## Results

Mean MOR availability in the subjects is presented in Figure 1. Mean weight loss at 3 months was 20.8 ± 5.6 kg, at 6 months was 25.7 ± 7.7 kg, at 12 months was 28.3 ± 12.1 kg, and at 24 months was 30.7 ± 15.1 kg. Postoperative weight development is shown in the Figure 2.

**Figure 1:**
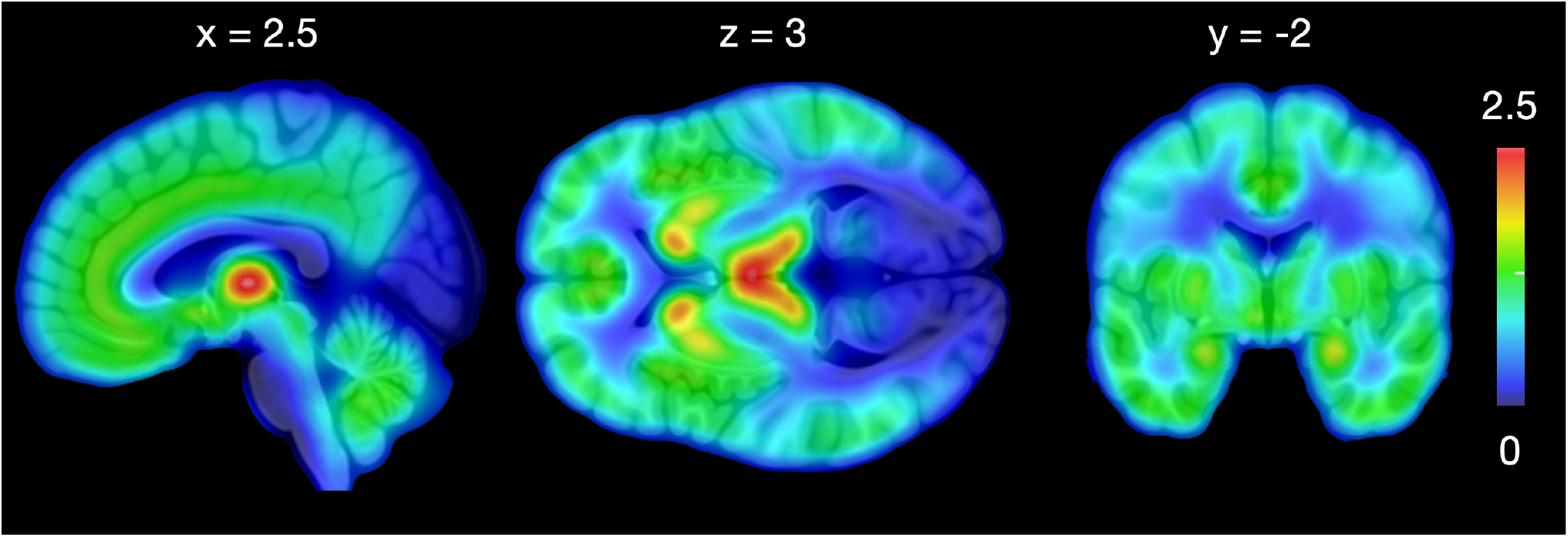
Mean [^11^C]carfentanil *BP*_ND_ in morbidly obese subjects before surgery.

**Figure 2:**
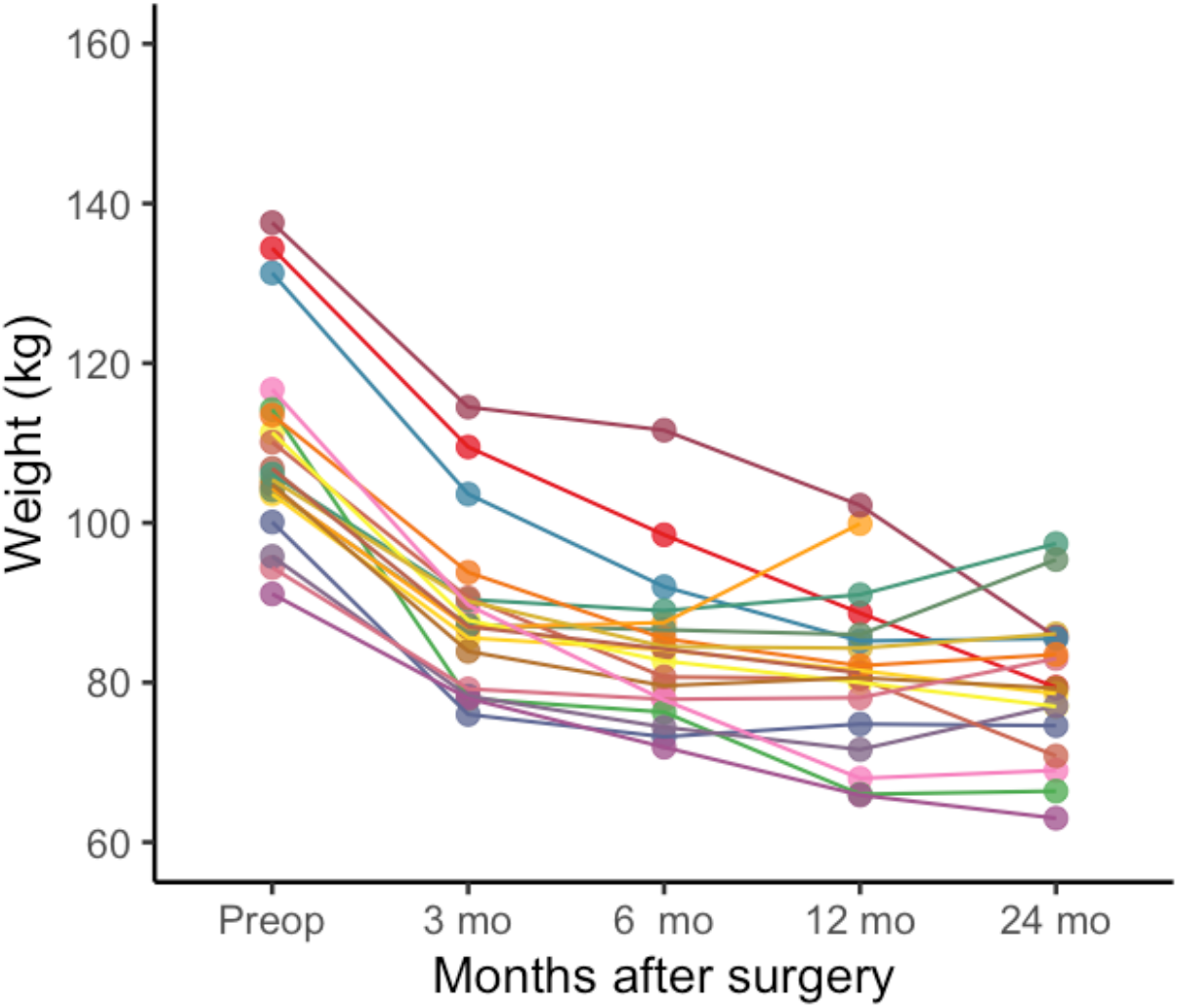
Weight development after bariatric surgery for each subject. Two subjects discontinued the study before 24 months follow-up visit.

Preoperative MOR availabilities were significantly associated with the subject weight in the amygdala (*r* = −0.54) (Figure 3), insula (*r* = −0.46), ventral striatum (*r* = −0.48) and putamen (*r* = −0.49) at 3 months. Significant association was also found in the amygdala at 6 months (*r* = −0.53) and at 12 months (*r* = −0.49) (Figure 3). Moreover, significant association was observed in the amygdala (*r* = −0.50) (Figure 2) and thalamus at 24 months (*r*s < −0.49).

**Figure 3:**
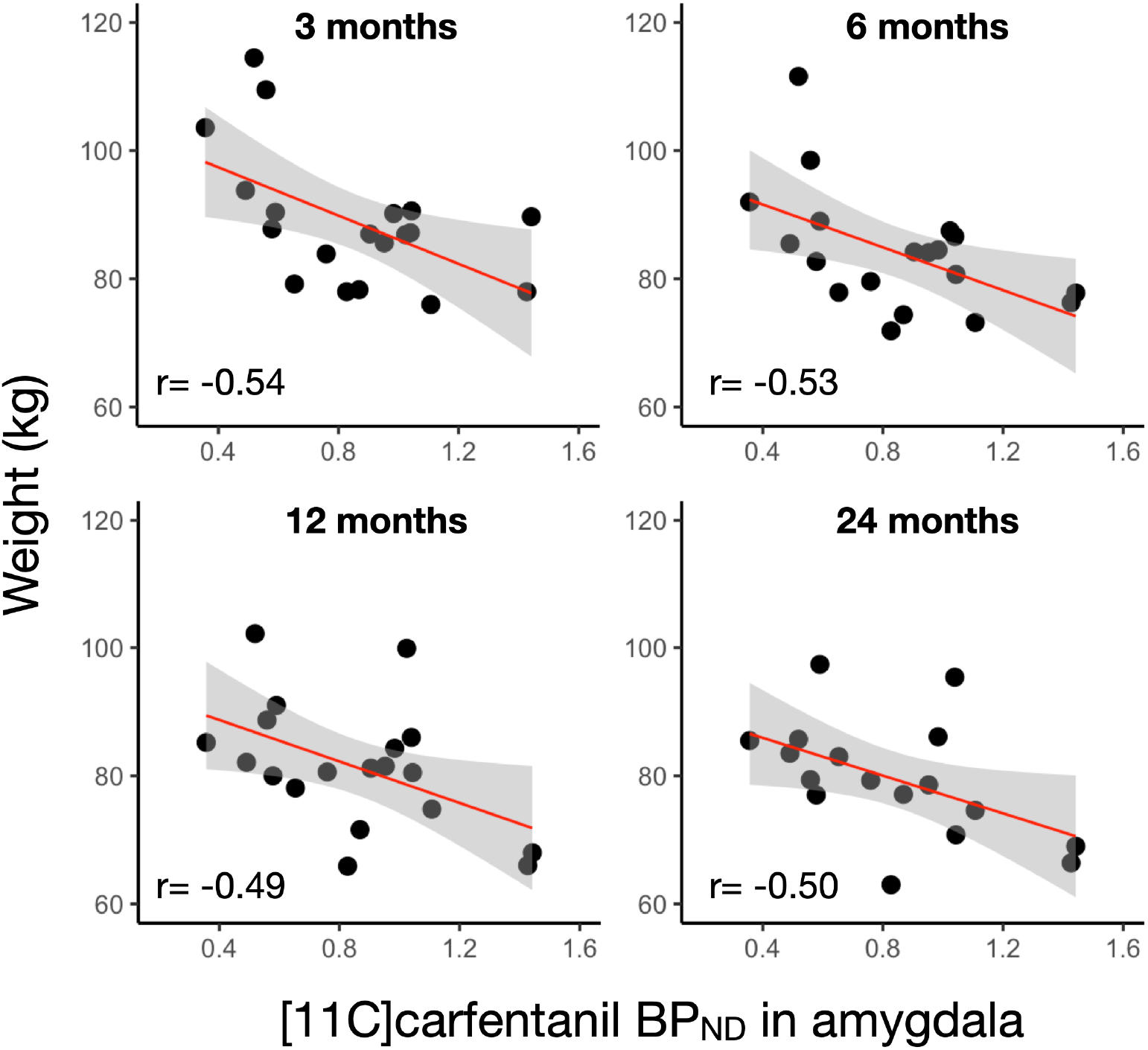
Correlations between preoperative [^11^C]carfentanil *BP*_ND_ in amygdala and subject weight at 3, 6, 12, and 24 months.

Preoperative weight did not correlate with MOR availabilities in any brain area. We did not find any significant correlation between preoperative D_2_R availability and subject weight in any brain area at any time point. No significant correlations between BDI-II and STAI scores and MOR and D_2_R availabilities in any brain area were observed. BDI-II and STAI scores did not predict weight loss at any time point.

## Discussion

Our main finding was that neuroimaging markers predict the weight loss outcome after bariatric surgery. MOR availability in the amygdala consistently predicted weight development throughout the 24-month follow-up period, even though MOR availability was not initially associated with preoperative weight. No associations were found for D_2_R. These results show that neuromolecular phenotypes may contribute to the outcome of weight loss after bariatric surgery, possibly providing novel predictive biomarkers for postoperative weight loss after bariatric surgery.

Obesity is expensive for the society, especially due to the obesity related comorbidities. Bariatric surgery reduces mean costs to the health service compared with usual care (Borisenko et al., 2018). However, a significant number of the patients experience weight regain (Monaco-Ferreira and Leandro-Merhi, 2017), which was also seen in our study (Figure 2). Determining patient characteristics leading to sustainable weight loss at long-term is important, but so far there have not been reliable markers. Some metabolic markers may predict weight regain after surgery (Abidi et al., 2019), also taste preference towards salty or sucrose-sweetened foods may contribute to some extent (Zhang et al., 2019; Smith et al., 2020). Our study is the first PET study to predict the outcome of bariatric surgery from neuroimaging markers, and only two small MRI studies exist (Holsen et al., 2018; Cerit et al., 2019). Our study shows that molecular organization of the brain’s reward circuit is an important determinant of the surgery-induced weight loss.

Bariatric surgery alleviates depressive and anxious symptoms (Dawes et al., 2016; Gill et al., 2019), yet psychiatric comorbidities are associated with weight gain following surgery (Mauro et al., 2019; Muller et al., 2019). Surgery has a more positive impact on the depressive disorders than anxiety disorders (de Zwaan et al., 2011), but preoperative symptoms also likely affect the results of the surgical methods. Preoperative depression is also associated with lower postoperative weight loss (Pedro et al., 2020). Although MOR availability is associated with depressive and anxious symptoms (Nummenmaa et al., 2020), we observed no association between depressive and anxiety symptoms and weight loss. This may be due to low statistical power for the questionnaire-based measures, as well as relative crudeness of questionnaires (in comparison with structural interviews such as MADRS).

Human PET studies have shown that feeding activates the endogenous opioid system (Tuulari et al., 2017), and consequently dysfunction of the endogenous opioid system is a key component underlying overeating, and thus a feasible target for pharmacological and behavioural interventions. Previous studies have investigated effects of bariatric surgery and following weight loss to separate receptor systems, showing mainly unaltered D_2_R availability and normalized MOR availability (Dunn et al., 2010; Steele et al., 2010; de Weijer et al., 2014; Karlsson et al., 2016), although one animal study has yielded contradictory findings (Hankir et al., 2016). Our study highlights the importance of MOR in the amygdala in predicting the weight management after the surgery. Opioidergic circuits in the amygdala are critical for emotions including fear and anxiety (Nummenmaa and Tuominen, 2018), but it is also one of the key regions in appetite control (Adhikari et al., 2015). MOR availability in amygdala is associated with subclinical depressive and anxiety symptoms (Nummenmaa et al., 2020), and it is likely that individual differences in MOR availability also in the amygdala may explain the differences in eating behaviour (Nummenmaa et al., 2018). It has also been shown that bariatric surgery can recover initially downregulated MOR in the amygdala of obese patients (Karlsson et al., 2016).

Our study has several limitations. Only female subjects were studied, and the results may not be generalizable to male subjects. Sample size was limited, possibly precluding establishing associations between MOR availabilities, weight development, and preoperative psychiatric symptoms. We followed the subjects only for two years as part of their standard clinical visits, but longer follow-up might have showed different trajectories. However, longer follow-up studies are planned in the future (Vreeken et al., 2019).

In summary, preoperative MOR availability in the amygdala predicts weight outcomes after bariatric surgery. Postoperative weight regain or primary weight loss failure may partially depend on dysfunctional opioid system. There is growing evidence that opioidergic system plays an important role in governing multitude of reward functions (Nummenmaa et al., 2018), and this study further confirmed its significance in the aspects of feeding (Tuulari et al., 2017). Downregulation of the MOR system can be reversed by surgical (Karlsson et al., 2016) and non-surgical weight loss (Burghardt et al., 2015). The present study extended these findings by establishing the role of MOR in long-term weight maintenance. Future prospective studies should address whether MOR availability is also predicative of weight gain in normal-weight subjects, and whether it predicts weight loss success by conventional dieting-based approaches.

## Data Availability

Due to the nature of this research, participants of this study did not agree for their data to be shared publicly, so supporting data is not available.

## Acknowledgments

The study was conducted within the Finnish Centre of Excellence in Cardiovascular and Metabolic Diseases supported by the Academy of Finland (grants #251125, #121031, #304385), Sigrid Juselius Foundation, University of Turku, Turku University Hospital and Åbo Akademi University. HKK was supported by personal grants from The Finnish Diabetes Research Foundation and The National Graduate School of Clinical Investigation. The funders had no role in study design, data collection and analysis, decision to publish, or preparation of the manuscript. The authors thank the staff of the Turku PET Centre for assistance in the PET imaging. Special thanks goes to research nurse Mia Koutu.

